# Rapid detection of SARS-CoV-2 variants of concern identifying a cluster of B.1.1.28/P.1 variant in British Columbia, Canada

**DOI:** 10.1101/2021.03.04.21252928

**Authors:** Nancy Matic, Christopher F. Lowe, Gordon Ritchie, Aleksandra Stefanovic, Tanya Lawson, Willson Jang, Matthew Young, Winnie Dong, Zabrina L. Brumme, Chanson J. Brumme, Victor Leung, Marc G. Romney

## Abstract

Using a real-time RT-PCR-based algorithm to detect SARS-CoV-2 variants of concern, we rapidly identified 77 variants (57-B.1.1.7, 7-B.1.351, and 13-B.1.1.28/P.1). This protocol enabled our laboratory to screen all SARS-CoV-2 positive samples for variants, and identified a cluster of B.1.1.28/P.1 cases, a variant not previously known to circulate in British Columbia.

## Introduction

A robust surveillance system for early identification of SARS-CoV-2 variants of concern (VOC) is of critical public health importance. VOC have demonstrated *in vitro* evasion of antibody neutralization by the K417N/T, E484K, and N501Y substitutions in the spike protein(1–3), and have displayed potential for enhanced transmission due to mutations within the spike receptor binding domain(4,5). Surveillance data from the United Kingdom demonstrated a rapid increase in SARS-CoV-2 cases beginning in September 2020, subsequently attributed to the B.1.1.7 variant(6). This variant has since become the predominant strain in several European countries, and its prevalence is increasing worldwide(7). The B.1.1.7 variant demonstrated dropout of the S gene on a commercial SARS-CoV-2 real-time RT-PCR assay in the UK due to a deletion mutation within the spike protein (69/70), and subsequently proposed as a surveillance strategy by the European Centre for Disease Prevention and Control(8).

The B.1.1.28/P.1 variant is an emerging VOC with cases first documented in Japan and Brazil. Within months, it has become the predominant strain in certain regions of Brazil, and has now been detected across several continents, although relatively few cases have been detected in Canada and the United States to date. Re-infection in patients with pre-existing SARS-CoV-2 immunity has been described, raising concerns about resurgence(9). Unlike B.1.1.7, the B.1.1.28/P.1 variant does not possess the 69/70 deletion mutation, highlighting the need for a versatile and comprehensive VOC surveillance strategy capable of detecting multiple mutations.

Given the potential for VOC to enhance transmission and mortality, and possibly evade natural and/or vaccine-induced immune responses, it is critical to identify VOC cases and monitor their prevalence. Though whole-genome sequencing (WGS) of random subsets of SARS-CoV-2 samples is a core surveillance strategy, this laborious and costly strategy is currently limited by lack of scalability and prolonged turnaround time. We propose a rapid VOC surveillance strategy using multiple real-time RT-PCR assays designed to detect key mutations among all specimens testing positive for SARS-CoV-2 in a clinical diagnostic laboratory.

## Methods

From January 25–March 1, 2021, nasopharyngeal swabs and saliva/mouth rinse samples with SARS-CoV-2 detected at any cycle threshold (Ct) value (reported as ‘positive’ or ‘indeterminate’) in our clinical virology laboratory were subsequently tested for VOC. SARS-CoV-2 detection was performed using the LightMix® ModularDx SARS-CoV (COVID19) E-gene assay (TIB Molbiol, Berlin, Germany), with use of the MagNA Pure Compact or MagNA Pure 96 and LightCycler 480, or with the cobas® SARS-CoV-2 Test (Roche Molecular Diagnostics, Laval, QC) on the cobas® 6800. VOC were detected with the VirSNiP SARS-CoV-2 Mutation Assays for Strain Surveillance (TIB Molbiol), targeting specific spike protein variations (N501Y, delHV69/70, K417N, E484K, V1176F). Samples were first screened for N501Y, and if detected, subsequent targets were re-tested to discriminate between the most prevalent VOC within our community (B.1.1.7-delHV69/70 and B.1.351-K417N) and newly emerging VOC (B.1.1.28/P.1-V1176F).

WGS was performed on the MinION (Oxford Nanopore Technologies) using the ARTIC nCOV-2019 sequencing protocol V.1 (J. Quick) using V3 primers or Illumina MiSeq with a modified ARTIC nCOV-2019 protocol. Accurate basecalling of MinION was performed using guppy 3.1.5, and FASTQ files analyzed on the bugseq.com platform using pangoLEARN. Illumina sequence data were analyzed with the in-house bioinformatics pipeline MiCall(10).

This study was approved by the Providence Health Care/University of British Columbia and Simon Fraser University Research Ethics Boards (H20-01055).

## Results

During the study period, 31,833 clinical samples were tested for SARS-CoV-2, with 2,618 instances of SARS-CoV-2 detected at any Ct value. Of these, 2,430 (92.8%) underwent subsequent testing for the three major VOC categories (B.1.1.7; B.1.351; B1.1.28/P.1). Note that 1.6% (38/2,430) of the samples failed to amplify using the N501Y assay, of which 71% (27/38) were reported as ‘indeterminate’ for SARS-CoV-2, reflecting late Ct values and presumably low viral loads. From the remaining 2,392 samples, 77 VOC were identified (57-B.1.1.7, 7-B.1.351, and 13-B.1.1.28/P.1). Over the course of the study, VOC detection among diagnostic samples rapidly increased (Figure). Notably, identified VOC included a large cluster of the B.1.1.28/P.1 variant not previously identified in British Columbia. All B.1.1.28/P.1 variants were initially suspected from the K417N assay, for which PCR products were identified at a lower melting temperature than expected. All the suspected B.1.1.28/P.1 variants were positive when re-screened using the V1176F target. The first presumptive B.1.1.28/P.1 variant identified was confirmed by WGS, detecting the following mutations in the S gene (characteristic of B.1.1.28/P.1): L18F, T20N, P26S, D138Y, R190S, K417T, E484K, N501Y, D614G, H655Y, T1027I, and V1176F. Sequencing also confirmed B.1.1.7 and B.1.351 samples detected by our PCR-based strategy.

**Figure.**
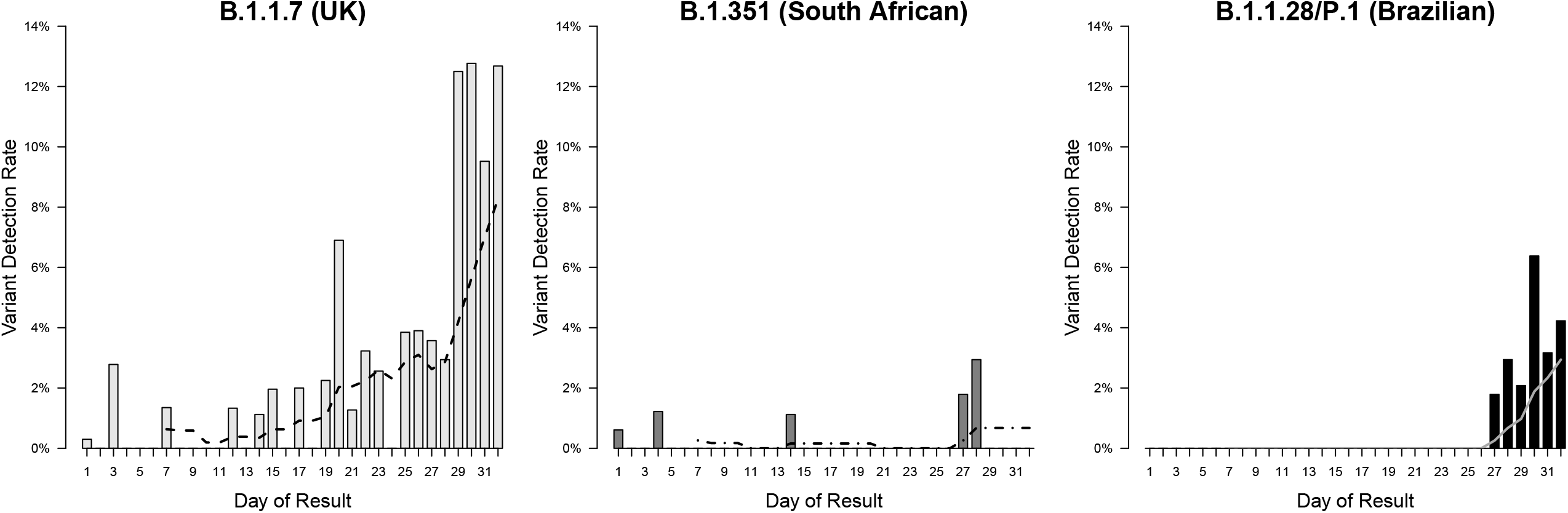
Detection rate of SARS-CoV-2 variants of concern by day of result for the study period of January 25 to March 1, 2021, with 7-day moving average

## Discussion

Timely identification is critical to identify emerging SARS-CoV-2 VOC that may be more transmissible, pathogenic and/or evade both natural and vaccine-induced immunity. The implementation of a PCR-based algorithm to detect VOC has enabled our laboratory to rapidly detect new variants that are in the very early stages of community transmission. Critically, our protocol enabled detection of VOC within 24 hours of COVID-19 diagnosis, a marked advantage over sequencing-based surveillance strategies. Over the study period, VOC positivity rate was 3.2%, but detection rates increased markedly over time, as might be expected with exponential growth observed in other countries.

Although B.1.1.28/P.1 had not previously been reported in our region, presence of this variant was suspected upon observation of K417N PCR products with a lower melting temperature than wild-type, suggestive of a non-K417N substitution such as the K417T mutation in the B.1.1.28/P.1 variant(11). Follow-up testing of these samples using the V1176F target supported the presumptive identification of B.1.1.28/P.1, which was subsequently confirmed by WGS. By testing all positive SARS-CoV-2 samples for VOC, it enabled the rapid detection of a discrete, new B.1.1.28/P.1 cluster.

Although the primary modality of VOC surveillance has been through WGS, universal sequencing of SARS-CoV-2 positive specimens is limited by both laboratory and bioinformatics capacity(12). Due to the volume of VOC testing required, high complexity and cost of WGS and limited laboratory capacity, turnaround time for VOC by WGS may be on the order of days to weeks. With the onset of the COVID-19 pandemic, molecular diagnostics (i.e., PCR) have been increasingly adopted by laboratories to promptly identify SARS-CoV-2 cases, with infrastructure established for this modality of testing. A PCR-based algorithm for the molecular detection of VOC could be rapidly adopted to provide almost real-time resulting of VOC to inform infection prevention and control and public health measures(7,13,14).

While the most prevalent VOC worldwide harbour the N501Y spike mutation, it is not present in all variants(15). A PCR-based algorithm for identifying VOC that uses N501Y as the initial screening target, such as in this study, must acknowledge this limitation. Given the rapid emergence of new variants, ongoing surveillance is key, and any laboratory considering a PCR-based algorithm would need to adapt the algorithm as VOC prevalence changes. Nevertheless, the ability to rapidly leverage existing molecular infrastructure established during the COVID-19 pandemic to presumptively identify the most prevalent VOC within 24 hours would be an important tool to complement WGS performed at reference laboratories.

In summary, our implementation of a real-time RT-PCR-based algorithm enabled identification of the most common VOC to date (B.1.1.7, B.1.351, and B.1.1.28/P.1) within 24 hours. This methodology would allow laboratories to perform VOC testing on all positive SARS-CoV-2 samples, and enhance VOC surveillance capacity to identify cases and support decision making for interrupting transmission.

## Data Availability

Data available on request from the authors

## Acknowledgements

We are grateful to our medical laboratory technologists who are highly committed to patient care and laboratory quality improvement. We would also like to acknowledge contributions from John Harding and Althea Hayden [Vancouver Coastal Health (VCH) Public Health], and the BCCDC Public Health Laboratory.

This work was supported by COVID-19 rapid response grants from GenomeBC (COV-115 to ZLB, CFL; COV-033 to CJB), an Exceptional Opportunities Fund – COVID-19 award from the Canada Foundation for Innovation (CJB, CFL), a British Columbia Ministry of Health - Providence Health Care Research Institute COVID-19 Research Priorities Grant (CJB, CFL) and Public Health Agency of Canada COVID-19 Immunology Task Force COVID-19 Hot Spots Competition Grant (ZLB, MGR). GR reports participation as a Roche Diagnostics Sequencing Advisory Panel member. ZLB holds a Scholar Award from the Michael Smith Foundation for Health Research.

